# Application of Principal Component Analysis to Heterogenous Fontan Registry Data Identifies Independent Contributing Factors to Decline

**DOI:** 10.1101/2024.07.11.24310309

**Authors:** Margaret R. Ferrari, Michal Schäfer, Kendall S. Hunter, Michael V. Di Maria

## Abstract

Single ventricle heart disease is a severe and life-threatening illness, and improvements in clinical outcomes of those with Fontan circulation have not yet yielded acceptable survival over the past two decades. Patients are at risk of developing a diverse variety of Fontan-associated comorbidities that ultimately requires heart transplant. Our observational cohort study goal was to determine if principal component analysis (PCA) applied to data collected from a substantial Fontan cohort can predict functional decline (N=140). Heterogeneous data broadly consisting of measures of cardiac and vascular function, exercise (VO_2max_), lymphatic biomarkers, and blood biomarkers were collected over 11 years at a single site; in that time, 16 events occurred that are considered here in a composite outcome measure. After standardization and PCA, principal components (PCs) representing >5% of total variance were thematically labeled based on their constituents and tested for association with the composite outcome. Our main findings suggest that the 6^th^ PC (PC6), representing 7.1% percent of the total variance in the set, is greatly influenced by blood serum biomarkers and superior vena cava flow, is a superior measure of proportional hazard compared to EF, and displayed the greatest accuracy for classifying Fontan patients as determined by AUC. In bivariate hazard analysis, we found that models combining systolic function (EF or PC5) and lymphatic dysfunction (PC6) were most predictive, with the former having the greatest AIC, and the latter having the highest c-statistic. Our findings support our hypothesis that a multifactorial model must be considered to improve prognosis in the Fontan population.

## Introduction

Patients born with single ventricle heart disease (SVD), a severe and rare congenital heart defect (CHD), are subjected to three palliative surgeries that culminate in the Fontan circulation [1]. While staged palliation addresses the primary concerns of obstructed systemic blood flow and cyanosis in a condition like hypoplastic left heart syndrome, a range of Fontan associated comorbidities, including lymphatic, liver, and cardiac damage, are often apparent in adolescence [2]. Because morbidity and mortality after Fontan surgery remain unacceptably high [2], new approaches to predict patient decline are sorely needed. The goal of this work is to develop a new prognostic model for patients with SVD and explore machine learning methods as a tool of risk stratification in the Fontan population.

Principal component analysis (PCA) is a data reduction technique that is often applied to large data sets in research [3–7], although it has not yet been applied to the Fontan population outside of waveform analysis [8, 9]. Examples where PCA has been utilized to find novel associations in large datasets that would have not have been amenable to more conventional statistical approaches include: Scientists have used PCA to identify patterns of inflammatory and adhesion molecules that contribute to muscle weakness acquired in the intensive care unit [4]. A similar study in a population of adults used PCA to identify inflammatory markers that precede major adverse cardiovascular events (MACE) following heart attack and found that the PC influenced by interleukin-6 and interleukin-8 was a better predictor of MACE at one year than univariate cytokine measures [6]. Additionally, PCA has recognized lymphocyte-monocyte-neutrophil indices that contribute to disease severity in several cohorts of COVID-19 patients [5]. The aforementioned studies support our hypothesis that a PCA approach may be necessary to understand and predict outcomes in heterogenous disease states like the Fontan population, where a very large number of potential predictor variables exist that stem from anatomic, surgical, imaging, laboratory, functional testing domains.

We have previously applied PCA to characterize non-pulsatile cavopulmonary flow waveforms [8, 9]; here we expand that approach to include heterogenous biomarkers. Another benefit of using a PCA approach in clinical data analysis is inclusion of correlative/colinear parameters, of which many statistical outcomes models prohibit [5]. Our primary objective in this study was to assess our previously defined novel waveform measures and other clinical parameters in a heterogenous PCA approach, all in support of the overall hypothesis that machine-learning extracted PCs will delineate patients with Fontan-associated comorbidities and reveal parameters that indicate circulatory failure in patients with a Fontan circulation. More specifically, a machine-learning extracted PC, which will consist of a pattern of abnormalities in multiple of cardiac and non-cardiac test results, previously unrecognized as an important preditor of outcomes, will be associated with a composite outcome of Fontan failure.

## Methods

One-hundred and forty SVD patients that underwent cardiac MRI (cMRI) at Children’s Hospital Colorado between July 2011 and August 2022 were included in this retrospective cohort study, permitted by the Colorado Multiple Institutional Review Board as a portion of Fontan at Altitude Registry for Outcomes (FAROUT). All patients cared for in the Fontan Multidisciplinary Clinic at the Children’s Hospital Colorado have undergone surveillance testing for end-organ damage and Fontan-associated comorbidities by way of a clinical practice guideline since 2016 and were included. The FAROUT registry was queried and abstracted data was used as a foundation for a study database, in addition to our single site venous flow patterns [10]. Variables collected are shown in Table I. For the purposes of survival analysis, study subject status was evaluated as of December 1, 2022, and a composite outcome was defined as the development of plastic bronchitis (PB, n=1), protein-losing enteropathy (PLE, n=2), referral to transplant (RTT, n=9), received a transplant (n=4), or death (n=0) from the time of cMRI to time to follow up.

**Table I.** Measures used in heterogenous PCA, including novel single vessel waveform data, hemodynamic, global cardiovascular, blood, kidney, liver, and respiratory biomarkers, and their mean or median and corresponding standard deviation or interquartile range.

| Measure | Mean or Median | St. dev or IQR |
| --- | --- | --- |
| SVC Single PC1 Scores | -0.522 | -2.87 - 3.12 |
| SVC Single PC2 Scores | -0.478 | -0.870 - 0.0638 |
| IVC Single PC1 Scores | -0.172 | -3.21 - 3.69 |
| IVC Single PC2 Scores | 0.21 | -1.53 - 1.35 |
| LPA Single PC1 Score | 0.397 | -3.01 - 2.07 |
| LPA Single PC2 Score | 0.181 | -0.656 - 1.24 |
| EF | 47.9 | 8.11 |
| EDVi | 93.3 | 27 |
| ESVi | 49.8 | 20.5 |
| SVi | 43.5 | 10.4 |
| CO | 4.43 | 1.43 |
| CI | 3.41 | 1.04 |
| BSA | 1.32 | 0.415 |
| BNP max | 30 | 15 - 61.8 |
| AST | 45 | 38 - 57 |
| VVCR | 0.963 | 0.3 |
| Lowest SaO2 | 86.4 | 5.56 |
| SVC Mean | 13 | 11 - 14 |
| mPAP | 12 | 10 - 13 |
| Alb | 4.54 | 0.685 |
| Platelets | 197 | 65.2 |
| Alk Phos | 158 | 80.3 |
| Total Protein | 7.53 | 1.1 |
| Creatanine | 0.61 | 0.49 - 0.763 |
| BUN | 14 | 12 - 17 |
| Cystatin C | 0.86 | 0.755 - 0.96 |

### cMRI Acquisition

Phase images and corresponding magnitude images of the superior vena cava (SVC), inferior vena cava (IVC), and left pulmonary artery (LPA) were obtained using a PC-MRI, ECG gated sequence as previously described [11, 12] by applying a 1.5 or 3.0 Magnetom Avanto (Siemens Medical Solutions, Erlangen, Germany) or Ingenia (Philips Medical System, Best, Netherlands) Tesla magnet using a phased-array body surface coil. A free breathing PC-MRI sequence was used under the following conditions: time to repetition, 14-28 milliseconds/25-40 cardiac phases; time to echo, 2.2-3.5 milliseconds; matrix, 160 x 256; flip angle, 25 degrees; 100% k space sampling; cross-sectional pixel resolution, 0.82 x 0.82 mm2 and 1.56 x 1.56 mm2; slice thickness, 5 millimeters. Heart rate dependent, PC-MRI acquisition varied 2-3 minutes for each vessel. Aliasing was accommodated for using the following velocity-encoding values: SVC and IVC, 75-100 cm/second; LPA, 50-100 cm/second. The AAo, SVC, and IVC images were acquired in axial cine stack and the LPA in vertical long axis, all orthogonal to flow.

### Flow Profile Analysis

Flow profile characteristics were assessed as described previously with slight modifications [9]. Flow waveforms were acquired by precise parallel segmentation of 2D phase-contrast image series in Circle CVi42 (Calgary, Canada). Flow data was captured for each patient at the SVC, IVC, and LPA and imported into MATLAB (Natick, MA). Each waveform was normalized by dividing each flow point by patient BSA to minimize size effect on the raw data. Data was interpolated using cubic spline interpolation to 40 points, guaranteeing size-matched array lengths for further analyses. Three data matrices were created containing single-site flow data and the size of each data matrix varied and is as follows: SVC (124 x 40), IVC (132 x 40), LPA (125 x 40).

### Clinical Biomarkers

Global cardiovascular and ventricular indicators (EF, CO, CI, EDVi, ESVi, SVi) have long been established as the benchmark for Fontan patient status [8, 13–15] and therefore were included in the analysis for validation purposes. VVCR, mean catheterization pressures (mPAP and SVC mean pressure), VO_2max_, BNP_max_ and O_2_ saturations were also included in the outcomes analysis, all of which have been independently linked to Fontan circulation health and outcomes [16–19]. We have previously determined that biomarkers indicative of lymphatic function and PLE, specifically aspartate aminotransferase, alkaline phosphatase, cystatin-c and creatinine were associated with caval flow patterns [9], strongly indicating that these parameters may identify, or be a predictive of, which Fontan patients will experience circulatory failure. Similarly, biomarkers such as albumin, total protein, blood urea nitrogen and platelet count were included due to previous reports relating these to Fontan patient cyanosis and pulmonary blood flow [20, 21].

### Principal Component Analysis

PCA requires that the input data matrix has a value assigned to each position, and therefore after the exported registry was read into MATLAB, patients with more than five measures missing were removed from analysis. This was done in an effort to maintain a missing data rate of less than 5% [22], and the resulting clinical parameters and demographic information can be found in Table I. Patients that had missing values less than or equal to five were replaced with the column median, as a value is required for each position in the input matrix. Columns were normalized by subtracting the column mean from each sample and dividing by the corresponding column standard deviation, and the resulting matrix was the input for PCA. Scree plots were created to determine PCs representing greater than 5% of total variance. Interpretation of which clinical parameters had the greatest influence on each PC were graphically determined by visualizing PC eigenvectors.

### Statistics

Statistical analyses were performed in GraphPad Prism and began by determining the univariate Cox hazard ratio (HR) for PCs 1-10 and EF, a measure of systolic function that serves as a benchmark diagnostic for patients with a Fontan circulation [23]. Akaike’s information criterion (AIC) and the c-statistic were also gathered, and measures with the greatest c-statistic were used to create receiver operating characteristic (ROC) curves and determine the area under the curve (AUC) and Youden’s index, defined as (sensitivity(x) – specificity(x)) −1. The optimum sensitivity, specificity and clinical threshold for grouping was found and used to defined groups for Kaplan-Meier survival analysis. The Mantel-Cox log-rank test was used to determine if a significant difference existed between Kaplan-Meier curves. Univariate parameters with the greatest c-statistics were used, up to two parameters at a time, for multivariate (bivariate) regression analysis and AIC was used to compare univariate and multivariate predictive models.

## Results

### Principal Component Analysis

The size of the original data matrix imported to MATLAB was 140 x 31 and was reduced to 115 x 31 after removal of patients missing greater than five measures. The remaining matrix had a 4.15% rate of missing data, and therefore fell within the 5% acceptable rate [22]. Columns of the matrix included scores for single site SVC, IVC, and LPA flow patterns, each representing a patient’s contribution to that PC’s waveform pattern, EF, EDVi, ESVi, SVi, CO, CI, BSA, BNP max, AST, VVCR, lowest SaO2, SVC mean pressure, mPAP, albumin, platelets, alkaline phosphatase, total protein, creatinine, BUN, and cystatin-C (Table I).

Following PCA, a scree plot was used to identify the percent each PC contributed to the overall variance in the original data set and can be seen in Figure 1. The first PC accounted for about 17.5% of the original data matrix variance, followed by approximately 11.5% for PC2, 8.5% for PC3, and subsequently decreased as the PC number increased (Figure 1). Each of the first 7 PCs accounted for more than 5% of the total variance, and together explained 70.3% of that variance; PCs up to PC10 (3.6% of total variance, 77.7% cumulative variance) were considered for survival analysis.

**Figure 1.**
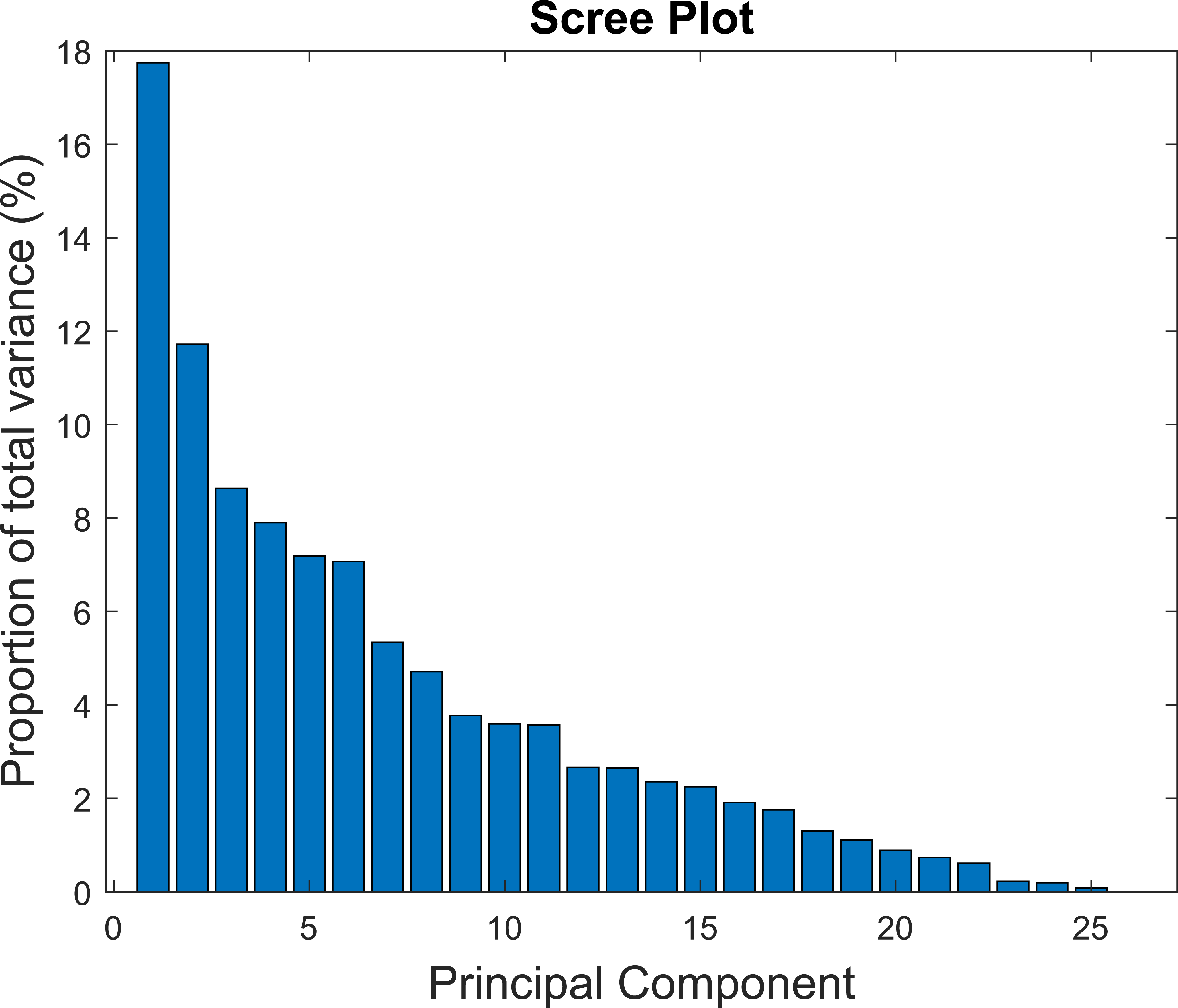
The scree pot displays each PC (x axis) and the percent variance it represents in the original data set (y axis).

Interpretation of the first two PCs was aided by the biplot displayed in Figure 2, where the blue lines represent the eigenvectors, or PC coefficients, and the length and direction represents that parameter’s influence on each PC. For example, further distance from the origin on the x-axis means greater contribution to PC1, therefore EDVi, ESVi, EF, and VVCR contributed greatest to PC1 (Figure 2). PC2 variance is explained by deviance from zero along the y-axis, and major influencing parameters include CO, SVi, LPA PC1 and IVC PC1 (Figure 2). The red data points represent the scores, or how each patient sample contributes to the PCs.

**Figure 2.**
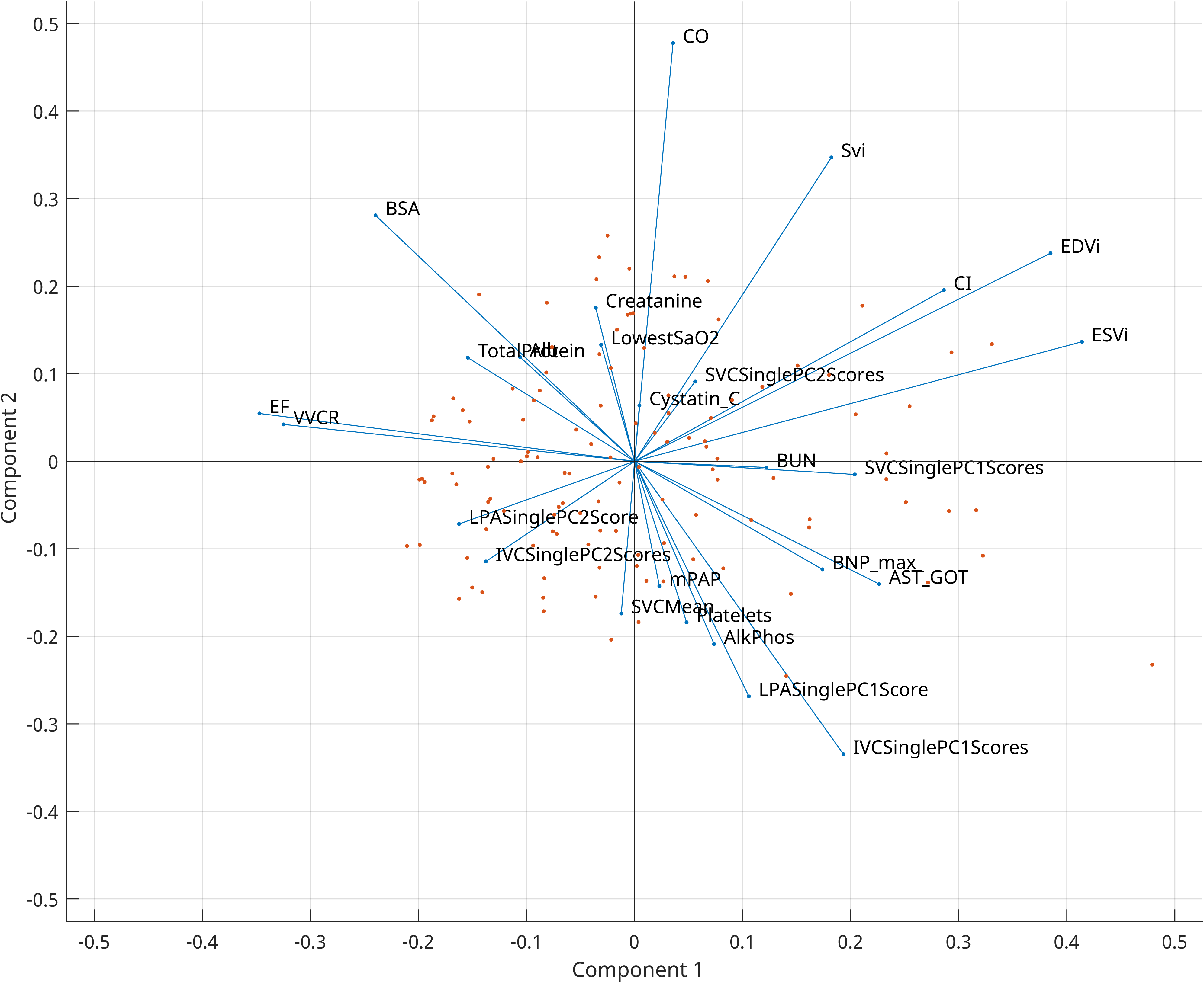
Principal component biplot that displays the scores returned from PCA, or each sample, in the original dimension and are represented by the red data points. The eigenvectors, or the coefficients, for each clinical parameter are displayed as the blue lines and the direction and length represent the influence each parameter has on PC1 (x axis) and PC2 (y axis).

The clinical implications of each PC were examined using bar graphs of the PC eigenvectors, where the clinical parameters (x axis) and their relative contributions to each PC (y axis) are displayed in Figure 3. The first PC was highly influenced by cardiac parameters, including EF, EDVi, ESVi, CI, VVCR, AST, SVC PC1 Scores, and IVC PC1 scores, which primarily describe cardiac function and the downstream effects in the Fontan circulation. The fourth PC was highly influenced by IVC and LPA waveform patterns in addition to cavopulmonary pressures and cystatin-C. PC5 was influenced by cardiac parameters representative of systolic function, such as EF, SVi, CI, and VVCR, and waveform patterns IVC PC2, SVC PC1 scores, and BUN. Albumin, alkaline phosphatase, total protein, BUN, BNP max, and SVC waveforms scores influenced PC 6.

**Figure 3.**
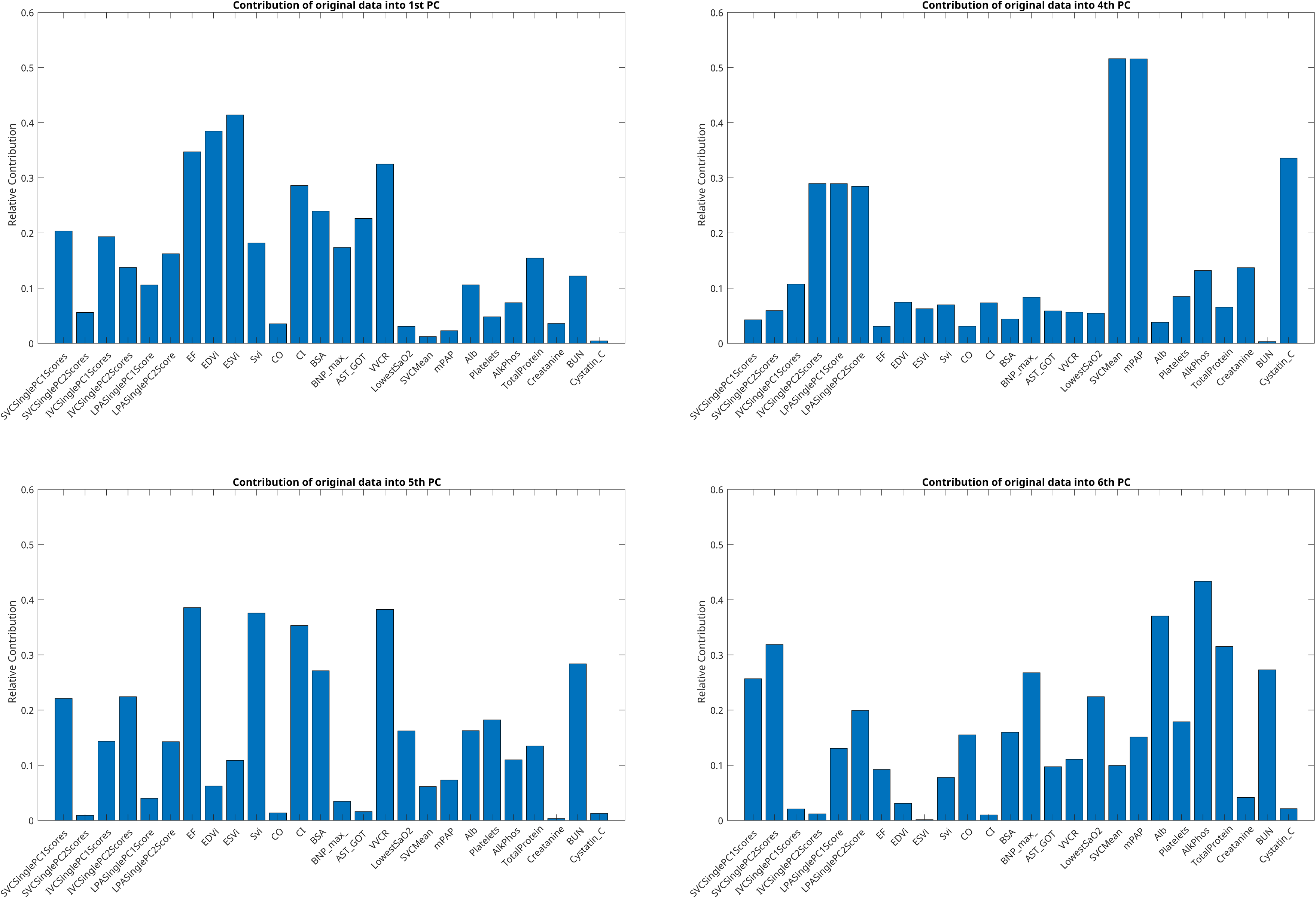
Bar graphs for PC1 and PCs 4-6 display each clinical parameter considered in PCA, or the column headers, and the amount (y axis, 0 up to 1) that parameter influences each PC.

### Survival Analysis

Univariate Cox proportional hazard ratio was determined for each PC and can be found in Table II. The single best predictor of which patient is at a greater hazard is PC6 (AIC=109), followed by the standard measure used for prediction in this population, EF (AIC=111) (Table II). PC1 and PC5 also performed well, with AICs of 113 and 115. The hazard ratio for EF and each PC is displayed in a forest plot in Figure 3, and the bars represent the 95% confidence intervals. If a parameter’s confidence interval crossed one, it was not statistically significant (Figure 4).

**Figure 4.**
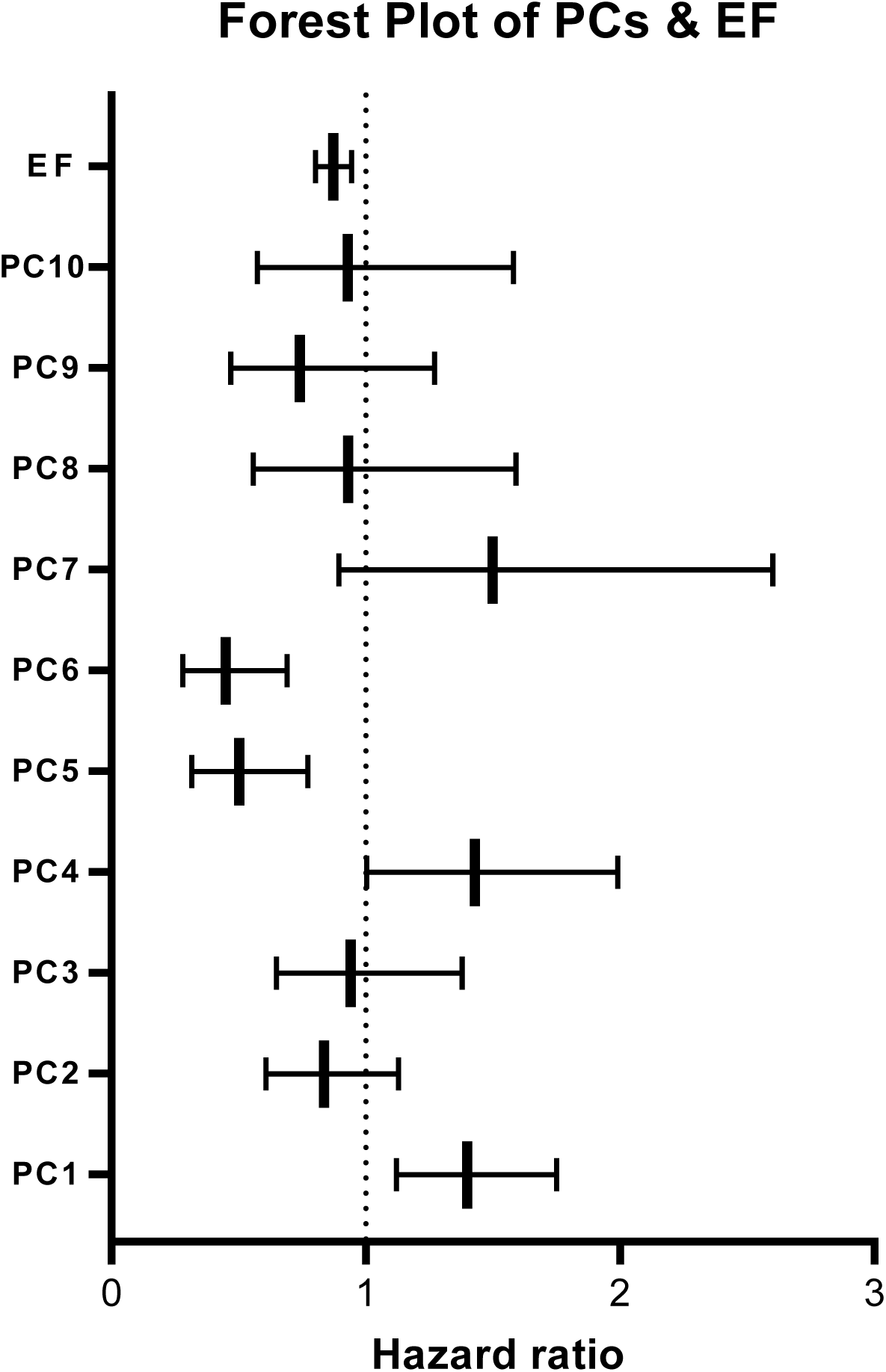
The forest plot displays the hazard ratio for each PC and EF and the corresponding 95% confidence intervals. Bars that cross 1, or the null hypothesis, represents no difference in hazards between patients that experienced an event versus those that did not.

**Table II.** Univariate cox regression hazard ratio for each PC and EF, the gold standard for systolic function and patient decline in the Fontan population, and the accompanying 95% confidence intervals, their p-value, c-statistic and AIC.

| **Variable** | **HR** | **95% CI** | **P value** | **c stat** | **AIC** |
| --- | --- | --- | --- | --- | --- |
| PC1 | 1.40 | 1.12 to 1.75 | 0.00280 | 0.705 | 115 |
| PC2 | 0.836 | 0.607 to 1.13 | 0.254 | 0.555 | 121 |
| PC3 | 0.941 | 0.649 to 1.38 | 0.753 | 0.453 | 123 |
| PC4 | 1.43 | 1.00 to 1.99 | 0.0404 | 0.619 | 119 |
| PC5 | 0.502 | 0.315 to 0.773 | 0.00250 | 0.718 | 113 |
| PC6 | 0.449 | 0.280 to 0.691 | 0.000500 | 0.759 | 109 |
| PC7 | 1.50 | 0.894 to 2.60 | 0.140 | 0.617 | 120.5 |
| PC8 | 0.931 | 0.558 to 1.59 | 0.793 | 0.587 | 123 |
| PC9 | 0.741 | 0.469 to 1.27 | 0.242 | 0.539 | 122 |
| PC10 | 0.929 | 0.574 to 1.58 | 0.777 | 0.519 | 123 |
| EF | 0.873 | 0.803 to 0.944 | 0.000900 | 0.726 | 111 |

Parameters with the greatest c-statistics were tested as classifiers of patients with Fontan decline using ROC curves, and PC6 returned the greatest AUC at 0.767, followed by PC5 (AUC=0.740), and EF (AUC=0.696) (Figure 5). The accompanying optimum sensitivity and specificity, determined using the greatest distance from the null hypothesis line or Youden’s index, was also found and shows that, while EF is highly specific (0.771) and therefore able to designate patients with Fontan failure correctly (low EF is almost always accompanied by SVD circulatory failure), its sensitivity is lacking at 0.643 (Figure 5). Sensitivity determines a classifier’s ability to label patients without Fontan decline correctly, and all PCs had the same, if not superior, sensitivity compared to EF (Figure 5). PC6 had the greatest sensitivity (0.786) with a reasonably balanced specificity at 0.686, suggesting it can rule healthy Fontan patients out as having circulatory failure, and PC1 had both optimum sensitivity and specificity at 0.714 (Figure 4). PC6 also displayed the greatest maximum effective biomarker, represented by Youden’s index of 0.471 (Figure 5).

**Figure 5.**
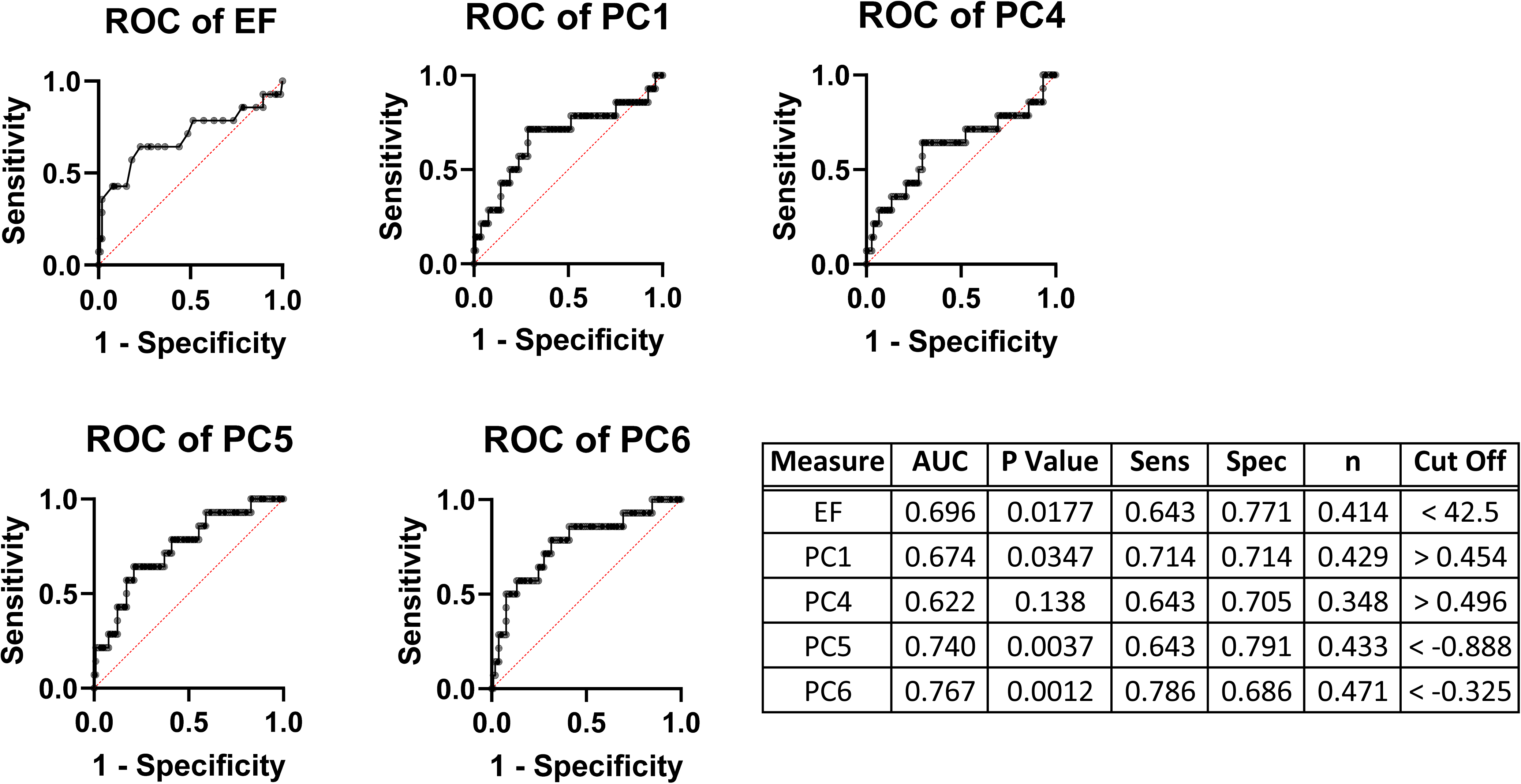
ROC curves for each PC that returned the greatest c-statistics and EF and the accompanying AUC, p-value, optimum sensitivity and specificity and the corresponding clinical cut off.

Grouping patients based on Youden’s index allows for Kaplan-Meier curve generation, displayed in Figure 6. Though EF was determined to have statistically significant differences in survival using the Mantel Cox log rank test (p=0.0006), PC1 and PC5 performed better with p values of 0.0003 and 0.0005 (Figure 6). PC6 also had highly significant differences in survival (p=0.0008), though it was not as significant as EF, and PC4 also displayed a significant difference in survival (p=0.07) (Figure 6).

**Figure 6.**
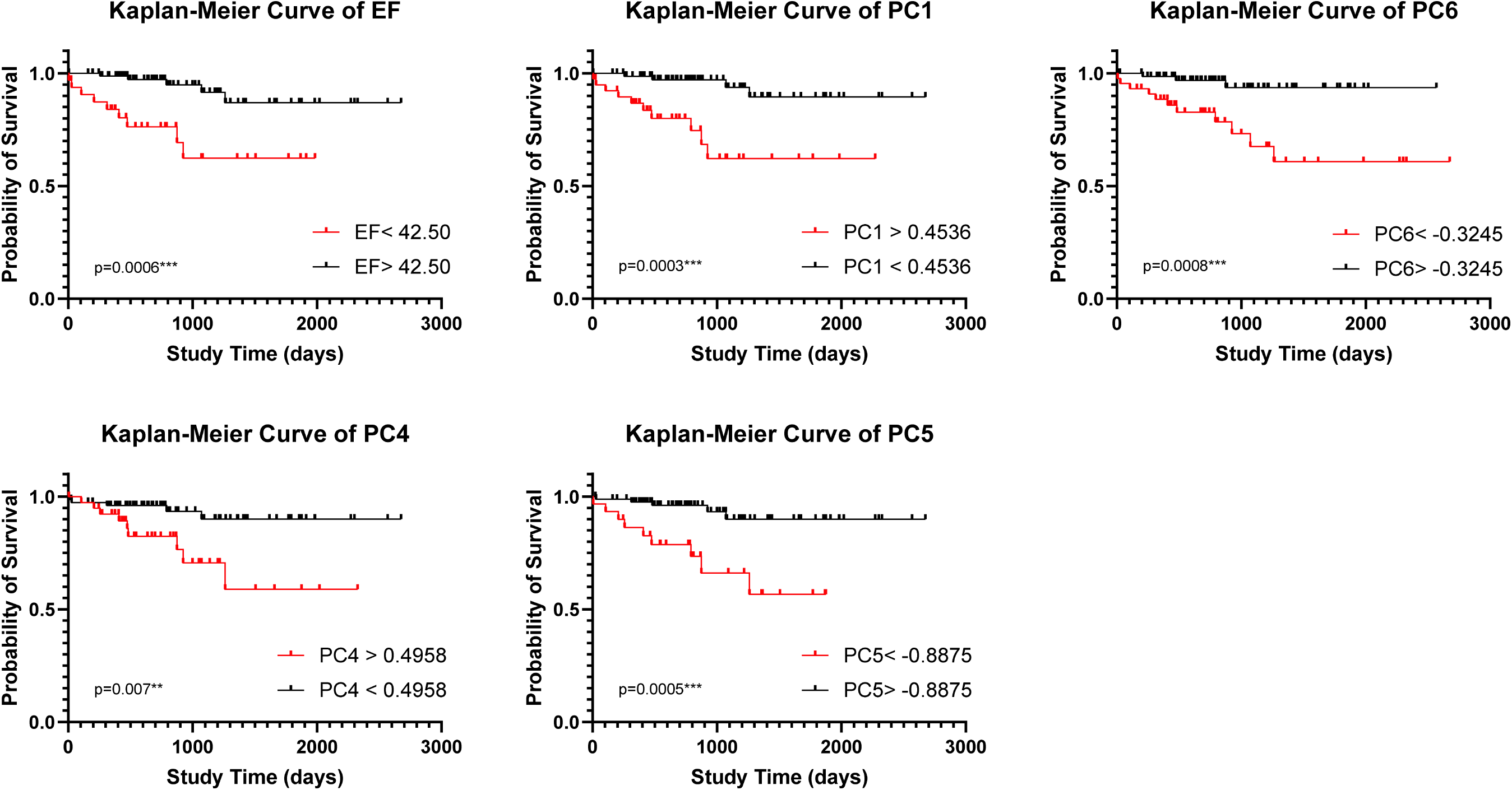
Kaplan-Meier curves for each PC that returned the greatest c-statistics and EF, and the p value returned from the log-rank Mantel Cox p-value. Groups were created using the Youden’s index defined cut-off values.

The inputs for multivariate Cox hazard regression analysis were EF, ESVi, and PC1 and PCs 4-6 and were chosen based on their univariate c-statistics. The models developed and each covariate’s HR estimate, 95% CI, p-value, c-statistic, and AIC are listed in Table III. The greatest AIC, and therefore predictive model, was model B (0.807, AIC=97) and included EF and PC6. However, model F, consisting of PC5 and PC6, has a greater c-statistic (0.845, AIC=103) and therefore is more probable to randomly identify a patient that experienced an event has a greater risk score than a patient that did not experience an event.

**Table III.** Multivariate models were developed, up to two measures at a time, for each PC and systolic measures EF and ESVi, the corresponding HR estimates, 95% CIs, p-values, c-statistic and AIC (used for model selection).

|  | **Covariates** | **HR Estimate** | **95% CI** | **p value** | **c stat** | **AIC** |
| --- | --- | --- | --- | --- | --- | --- |
| **A** | EF | 0.906 | 0.804 to 1.02 | 0.101 | 0.726 | 112 |
|  | ESVi | 1.02 | 0.979 to 1.06 | 0.413 |  |  |
| **B** | PC6 | 0.455 | 0.287 to 0.682 | 0.0003 | 0.807 | 97 |
|  | EF | 0.875 | 0.811 to 0.939 | 0.0003 |  |  |
| **C** | PC5 | 0.667 | 0.402 to 1.04 | 0.0931 | 0.738 | 110 |
|  | EF | 0.901 | 0.822 to 0.988 | 0.0247 |  |  |
| **D** | PC4 | 1.37 | 0.960 to 1.93 | 0.0765 | 0.756 | 110 |
|  | EF | 0.884 | 0.814 to 0.952 | 0.0019 |  |  |
| **E** | PC1 | 1.11 | 0.802 to 1.52 | 0.505 | 0.734 | 112 |
|  | EF | 0.895 | 0.802 to 0.994 | 0.0413 |  |  |
| **F** | PC5 | 0.574 | 0.372 to 0.841 | 0.0071 | 0.845 | 103 |
|  | PC6 | 0.461 | 0.279 to 0.721 | 0.0012 |  |  |
| **G** | PC4 | 1.395 | 0.982 to 1.95 | 0.0558 | 0.803 | 108 |
|  | PC6 | 0.445 | 0.271 to 0.695 | 0.0007 |  |  |
| **H** | PC4 | 1.448 | 1.01 to 2.06 | 0.0426 | 0.764 | 111 |
|  | PC5 | 0.501 | 0.313 to 0.771 | 0.0024 |  |  |
| **I** | PC1 | 1.322 | 1.06 to 1.65 | 0.0123 | 0.799 | 105 |
|  | PC6 | 0.516 | 0.327 to 0.766 | 0.0022 |  |  |
| **J** | PC1 | 1.595 | 1.22 to 2.11 | 0.0006 | 0.784 | 103 |
|  | PC5 | 0.482 | 0.313 to 0.717 | 0.0005 |  |  |
| **K** | PC1 | 1.404 | 1.12 to 1.75 | 0.0028 | 0.781 | 113 |
|  | PC4 | 1.462 | 1.01 to 2.10 | 0.0425 |  |  |

## Discussion

In this study, we explored univariate and multivariate associations between composite outcomes in a relatively large, single-center Fontan cohort, evaluating standard and ML-derived parameters both singly as well as through heterogeneous feature reduction (PCA). Through use of PCA, we determined PCs that appear to relate to specific features of Fontan decline, and that these features were significant univariate and multivariate predictors of a composite event.

PC6 was identified as a highly significant measure of hazard in our cohort and was the greatest univariate predictor of outcomes with an AIC of 109. The parameters that contributed the greatest to this PC have also been associated with development of lymphatic dysfunction, specifically PLE, in the Fontan population and include albumin, alkaline phosphatase, total protein, BUN and BNP max [23–25]. Cavopulmonary flow patterns were also found to influence PC6 and have previously been suspected as contributors to PLE [23]. PC6 was also the most accurate classifier of patients with Fontan decline from those without, which supports our understanding of PLE development and poorer prognosis in the Fontan population. It is also worth noting that only two patients (of 16) experienced a composite outcome of PLE. Additionally, with a sensitivity of 78.6% and specificity of 68.6%, this PC may be clinically useful in categorizing (or risk stratifying) patients. For example, after undergoing surveillance testing for end-organ damage and Fontan-related comorbidities, a patient could have their CMR-derived flow waveforms and other biomarkers examined here projected into a known feature set (heterogeneous PCs), after which these latter PCs would be used to classify the patient as either at-risk or not, based on the present analysis. Additional work, in terms of data collection and validation, as well as potentially longitudinal studies targeting causality, must be performed to identify the role parameters represented by PC6 play in the development of PLE, though this study provides a promising foundation for further research.

Survival was significantly predicted by all PCs identified as having a significant hazard ratio (1, 4, 5, 6), though two performed superior to EF, the standard measure of health and cardiac function in this population. PC1 performed better at prediction than EF alone and represented components of general cardiac function, including EF, EDVi, ESVi, VVCR, CI, and cavopulmonary flow measures. Our findings suggest that PCA is a supported method for inclusion of colinear parameters, and inclusion of such measures does in fact improve prediction of outcomes better than a single measure of ventricular function or multivariate testing. PC5 also improved prediction of outcomes compared to EF and was influenced by EF, SVi, CI, BSA, VVCR, BUN, SVC and LPA waveform patterns, most of which are affected by systolic ventricular function. However, little improvement was noted in a multivariate model that included EF and PC1 (AIC=112) and EF and PC5 (AIC=110) compared to the individual parameter’s AIC scores [EF (111), PC1 (115), and PC5 (109)] which may be due to redundant information (i.e. EF, and thus, systolic function, is now accounted for twice in the model). These results suggest that a PCA approach can improve outcomes prediction in the Fontan population and continue to support the hypothesis that machine-learning extracted PCs will clearly delineate SVD patients with Fontan-associated comorbidities to those without, in addition to our previously published hypothesis that a multivariable approach, in this case PCA, improves prediction of this heterogenous patient population with multiple organ systems in various stages of failure [9].

The best predictive model explored in this study, determined by AIC, is the multivariate Cox regression model B (Table III) including covariates EF and PC6. If PC6 is in fact linked to PLE, our findings suggest a combination of systolic ventricular function and measures indicative of lymphatic dysfunction may be an avenue for improved prognostication. This model, however, did not have the greatest c-statistic, which suggests it may not be suitable for ranking patients according to risk. Model F, including covariates PC5 and PC6, had the greatest c-statistic and is most suitable in determining which patients are at a higher risk.

The limitations of this study, as previously described [8], include those inherent to PCA. Linear data reduction does not consider non-linear reduction methods and, as the name suggests, compresses the original data for usability and is accompanied by a loss of, ideally insignificant, original data variance. Additionally, several patients were removed from analysis because PCA requires that the input matrix has no missing data. The cohort may contain a selection bias, because not all of our Fontan patients received a routine CMR examination in the past. Finally, machine learning methods thrive on large datasets, and while the final set used for PCA (N=115) is large for a pediatric population, clearly multicenter studies or learning networks that pool such data will offer even greater insights into disease progression. Despite limitations, this work has established that a heterogenous approach to PCA is beneficial to outcomes prediction in Fontan patients, and that our novel single site venous waveform patterns contribute to PCs predictive of decline.

### Conclusion

The goal of this study was to determine if a heterogenous PCA approach applied to the Fontan cohort can predict functional decline in this population. Our main findings suggest that PC6, which represented roughly 7% of the overall variance and is greatly influenced by blood serum biomarkers and SVC flow, is a superior measure of proportional hazard in this population compared to EF. We also found that PC6 displayed the greatest accuracy for classifying Fontan patients, as determined by AUC, and we identified two PCs that indeed predicted survival in this population better than EF. Our findings support our suspicions that a multifactorial model must be considered to improve prognosis in the Fontan population.

## Data Availability

The data that support the findings of this study are available from the corresponding author, KSH, upon reasonable request

## Acknowledgements

This research was supported by the Jayden DeLuca Foundation, NIH CTSA Grant UL1 TR002535, and the American Heart Association Children’s Heart Foundation Predoctoral Congenital Heart Defect Research Award 20PRE35260057 to MRF. The authors would like to additionally thank Dr Dunbar Ivy for his support of the dissertation work that led to this manuscript.

## Abbreviations

SVD: single ventricle disease
PLE: protein losing enteropathy
PB: plastic bronchitis
FALD: Fontan-associated liver disease
PC MRI: phase contrast magnetic resonance imaging
VVCR: ventricular vascular coupling ration
VO_2_: rate of oxygen consumption
AAo: ascending aorta
SVC: superior vena cava
IVC: inferior vena cava
LPA: left pulmonary artery
cMRI: cardiac MRI
TCPC: total cavopulmonary connection
PCA: principal component analysis
PCs: principal components
EF: ejection fraction
EDVi: end diastolic volume index
ESVi: end systolic volume index
CI: cardiac index
BNP: B-type natriuretic peptide
GGT: gamma-glutamyl transferase
AST: aspartate aminotransferase
SaO_2_: arterial oxygen saturation
FEV1: forced expiratory volume in one second
mSVCP: mean SVC pressure
mPAP: mean pulmonary artery pressure
HLHS: hypoplastic left heart syndrome
TA: tricuspid atresia
DORV: double outlet right ventricle
DILV: double inlet left ventricle
HRHS: hypoplastic right heart syndrome
TCPC: total cavopulmonary circuit
FVC: forced vital capacity
RV: residual volume
TLC: total lung capacity
Alk phos: alkaline phosphatase
BUN: blood urea nitrogen

